# Discordance Between Self-Reported and Lab-Measured A1C Among U.S. Adults with Diabetes: Findings from the National Health and Nutrition Examination Survey (2013-2020)

**DOI:** 10.1101/2023.05.10.23289782

**Authors:** Aneesh Kamath, Christopher C. Imes

**Author notes:** **Corresponding Author** Aneesh Kamath, 3500 Victoria St., 336 Victoria Building, Pittsburgh, PA 15261, USA.

## Abstract

**Aims:** To: 1) compare characteristics of those who report knowing their hemoglobin A1C (A1C) value versus those who do not; 2) determine the correlation and concordance between self-reported and lab-measured A1C; and 3) examine factors associated with a lab-measured A1C of ≤ 7%.

**Methods:** This was a cross-sectional secondary data analyses of the National Health and Nutrition Examination Survey from 2013-2020. Participants ≥ 20 years old who reported receiving a diabetes diagnosis were included.

**Results:** After proper sample weighting, twenty-two percent of participants reported not knowing their A1C value. Not knowing one’s A1C value was associated with identifying as a racial or ethnic group other than White, having a lower income, and having less formal education (P values < 0.5). Self-reported A1C was moderately correlated with lab-measured A1C (r = 0.62, P < 0.001). Higher self-reported A1C and identifying as Black or Mexican American were associated with lower odds of good glycemic control.

**Conclusions:** Many patients with diabetes did not know their A1C, and among those that did, the value was often inaccurate. Even when patients knew their A1C, the correlation between self-reported and lab-measured A1C was only moderate. Clinicians should evaluate and, if needed, enhance patient knowledge of A1C.

## 1. Introduction

Diabetes is a chronic health condition affecting over 34 million adults (including undiagnosed diabetes) in the United States (U.S.) and greatly increases risk of mortality [1]. In 2017, the estimated total cost of diabetes, including lost productivity, exceeded $327 billion [2]. The prevalence of diabetes has been increasing since 1988, while good glycemic control among patients with diabetes has been declining since 2010 [3,4]. Poor diabetes management is associated with nephropathy, neuropathy, retinopathy, and ketoacidosis [5-8]. Type 2 diabetes, the most common form of diabetes, is additionally associated with an increased risk of adverse cardiovascular outcomes [9]. Better glycemic control has been repeatedly shown to reduce risk of diabetes-related complications [10-12]. Hemoglobin A1C (A1C) has been recognized as an important measure of glycemic control [11,12]. An A1C of ≤ 7% is the goal for most adults with diabetes [13]. Patient knowledge and understanding of A1C are considered essential requirements for diabetes self-management [13-15].

In the U.S., individuals identifying as racial and ethnic minorities and those without health insurance are less likely to have their A1C levels tested [16]. Among those who have had their A1C tested, two recent small studies suggest that the majority of patients are aware of A1C as an important factor in diabetes management [17,18]. However, this awareness of A1C did not include how A1C correlates with average blood glucose levels [17,18]. Only one of the studies examined the concordance between perceived (i.e., self-reported) A1C and lab-measured A1C [17]. The study found that 79.8% of participants reported knowing their last A1C test result [17]. For those individuals who knew their last A1C test result, the perceived A1C was congruent with lab-measured A1C for 74.6% [17]. Unfortunately, the operational definition of congruence was not provided. However, an earlier study by Heisler and colleagues reported that 44% of patients knew their A1C value and only 25% could report the value correctly [19]. The study’s operational definition of correctly knowing one’s A1C value was being within 0.5% of the lower or upper boundary of the participant’s chosen A1C response category (< 7; 7–8; 8–9; 9–10; > 10; and don’t know). These three studies recruited participants from clinical practice settings. Thus, the findings may not be generalizable to all individuals with diabetes. Whereas, awareness and understanding of A1C varies, there is evidence that patients with a better understanding of their A1C have better diabetes self-management [19].

From an international perceptive, only Trivedi and colleagues have examined individuals’ knowledge of A1C testing and the concordance between self-reported and lab-measured A1C [15]. In that study, 48% of participants reported knowing their A1C, and 78.3% of those had self-reported values within 0.5% of their lab-measured A1C. Self-reported and lab-measured A1C were in high agreement, measured by the intraclass correlation coefficient. Participants whose self-reported and lab-measured A1C differed by more than 0.5% tended to have worse glycemic control.

While research on the agreement between self-reported A1C values in comparison to lab-measured A1C in the U.S. has been conducted [17-19], studies using a nationally representative sample are lacking. Additionally, studies have neither examined the concordance between self-reported and lab-measured A1C among racial and ethnic groups nor by important sociodemographic factors like sex and educational attainment. Therefore, the purpose of this secondary analysis of data from the National Health and Nutrition Examination Survey (NHANES) was to: 1) identify characteristics associated with knowing one’s A1C value versus not knowing; 2) determine the correlation and concordance between self-reported and lab-measured A1C; and 3) examine the factors associated with good glycemic control defined as a lab-measured A1C of ≤ 7%.

## 2. Methods

### 2.1 Sample

NHANES is a survey of residents of the U.S. that conducts health interviews and a medical examination which collects laboratory measures such as A1C [20,21]. We used NHANES data from 2013-2014, 2015-2016, and 2017-2020. NHANES data is publicly available and was approved by the Center for Health Statistics Research Ethics Review Board [22]. NHANES collected written informed consent from all adult participants and the data were anonymized. NHANES randomly selects participants in a manner such that each cycle is representative of the U.S. population as a whole [22]. NHANES methods also account for non-response bias and missing data [22]. Therefore, our results are generalizable to the U.S. population [22].

For the study, we included participants 20 years or older who reported receiving a diabetes diagnosis with laboratory measured A1C. Exclusion criteria was refusal to provide self-reported A1C.

### 2.2 Study variables

#### 2.2.1. Diabetes diagnosis

NHANES participants were asked if they had a diagnosis of diabetes.

#### 2.2.2. Self-reported A1C

NHANES interviewers asked participants who self-reported receiving a diabetes diagnosis if they had received an A1C test in a healthcare setting within the last year. Participants responded with the numerical value, “Don’t know”, “Refused”, or reported no A1C test.

#### 2.2.3. Lab-measured A1C

During the physical examination, fasting blood samples were obtained for A1C levels. Measures were obtained with appropriate protocols for quality and consistency, using an assay which could differentiate between glycated and non-glycated forms of A1C [23]. The percent of glycated hemoglobin was measured [24-26]. Given that a 0.5 change in A1C is considered statistically and clinically significant [27], the authors defined A1C concordance as the magnitude of difference between self-reported and lab-measured A1C as ≤ 0.5. Differences > 0.5 was defined as A1C discordance. This is consistent with the approaches used by Trivedi and colleagues [15].

#### 2.2.4. Sociodemographic information

Participants self-reported age. For all analyses and regression models, age was categorized as 20-39, 40-59, and 60 years and older [28]. Participant sex was self-reported as male or female. Race and ethnicity were self-identified and included Mexican American, non-Hispanic White, non-Hispanic Black, non-Hispanic Asian, other Hispanic, and other race, in accordance with the definitions provided by NHANES operational plan [20].

#### 2.2.5. Poverty-income ratio (PIR)

NHANES calculated PIR, which estimates income by adjusting for cost of living and family size. Consistent with other NHANES studies, the authors stratified the PIR as ≤ 130%, > 130% and ≤ 350%, and > 350% of the federal poverty level [29,30].

#### 2.2.6. Educational attainment

Participants self-reported their highest level of educational attainment.

#### 2.2.7. Clinician’s recommended target A1C

Individuals who self-reported diabetes were asked what their healthcare provider recommends as their target A1C [31]. Participants reported the value, having no target, or did not know.

### 2.3. Statistical Analyses

Data analyses were conducted using R (version 4.1.2) and packages survey (version 4.1.1), jtools (version 2.1.4), and irr (version 0.84.1) [32-36]. Proper survey weighting was used in all analyses to account for complex survey design. Weighting accounts for differing probability of selection due to oversampling certain populations and survey nonresponse [37].

Descriptive statistics are reported as unweighted count and weighted percentage for categorical variables, and mean and standard deviation (SD) for continuous variables. As a sensitivity analysis, the sample was stratified based on whether or not participants reported knowing their A1C. This method of analysis allows elucidation of differences between those two groups. Differences in characteristics between groups were determined using the student t-test for continuous variables and the Chi-Square test for categorical variables. All significance tests were two-sided and the significance level was 0.05. These analyses were adjusted for multiple comparisons using the Hochberg method [38].

For visualization of self-reported A1C values versus lab-measured A1C, a survey weighted scatterplot was created. Local polynomial regression was used to visualize the linear trend. The model was plotted on the scatterplot against the 45-degree line representing perfect equality between self-reported and lab-measured A1C. Additionally, a Bland and Altman plot was created to determine agreement between the two measures [39].

To determine correlations between self-reported A1C and lab-measured A1C, intraclass correlation coefficient (ICC) and Pearson’s correlation coefficient [40-41] were used. Correlations were covariate adjusted with a priori covariates of age, sex, race and ethnicity, PIR, educational attainment, and insurance. These covariates were selected from an examination of the literature and availability of data in NHANES [42-43]. P-values or confidence intervals (CIs) were calculated for all correlation coefficients using bootstrapping techniques with 100,000 iterations. Subgroup analyses by age, sex, race and ethnicity, PIR, educational attainment, and insurance were conducted. Additionally, Spearman (*ρ*) and the Lin (*ρ*_*C*_) correlation coefficients were created [44].

A1C concordance and discordance were calculated using survey-weighted 95% CIs, which provides a nationally generalizable estimate of concordance and discordance among participants with self-reported diabetes.

A logistic regression model was constructed with glycemic control, defined as lab-measured A1C ≤ 7% as the dependent variable and self-reported A1C as the independent variable, adjusted for the a priori covariates reported above. Odds ratios were reported using the estimate and 95% CI.

## 3. Results

Our sample was composed of 2,723 participants with self-reported diabetes (see figure 1). The sample consisted of mostly non-Hispanic White participants (n = 867, weighted percentage = 60.5%) and slightly over half were male (n = 1,443, 53.6%). The mean age was 60.4 years (SD = 13.3). With regard to income, 853 participants (24.4%) had income ≤ 130% of the federal poverty level while 599 participants (36.4%) had incomes > 350% of the federal poverty level. For educational attainment, most participants were either high school graduates or obtained a GED (n = 648, 27.0%) or had some college or AA degree (n = 818; 33.1%). The mean self-reported A1C was 7.2% (SD = 1.6). The mean of lab-measured A1C was 7.4% (SD = 1.7). With respect to clinician provided target A1C, 290 (9.4%) participants did not know, 659 (34.6%) had < 7% as their target A1C, and 732 (35.3%) had a target A1C of < 6%.

**Figure 1.**
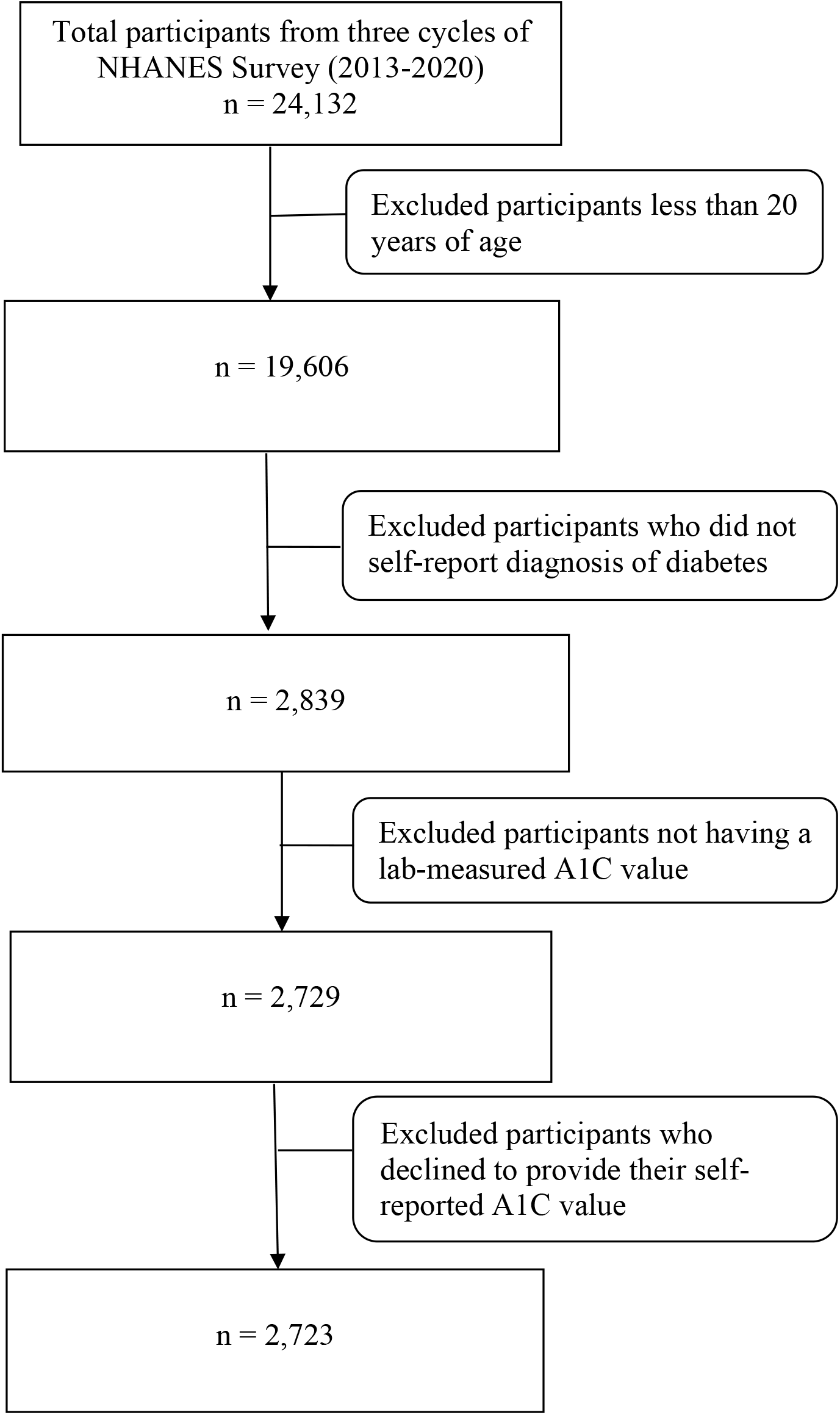
Flowsheet showing total participants for this study.

Twenty-two percent of participants did not know their A1C value from their last A1C test. Not knowing one’s A1C value was associated with identifying as a racial or ethnic group other than non-Hispanic White, having a lower PIR, and having a less formal education (P < 0.05 for all measures). There were no statistically significant differences with respect to age, sex, and health insurance status among participants who knew their A1C value and those who did not. Those who did not know their A1C were more likely to not know or have an unspecified clinician-provided target A1C goal (P < 0.001). See table 1 for details.

**Table 1.**
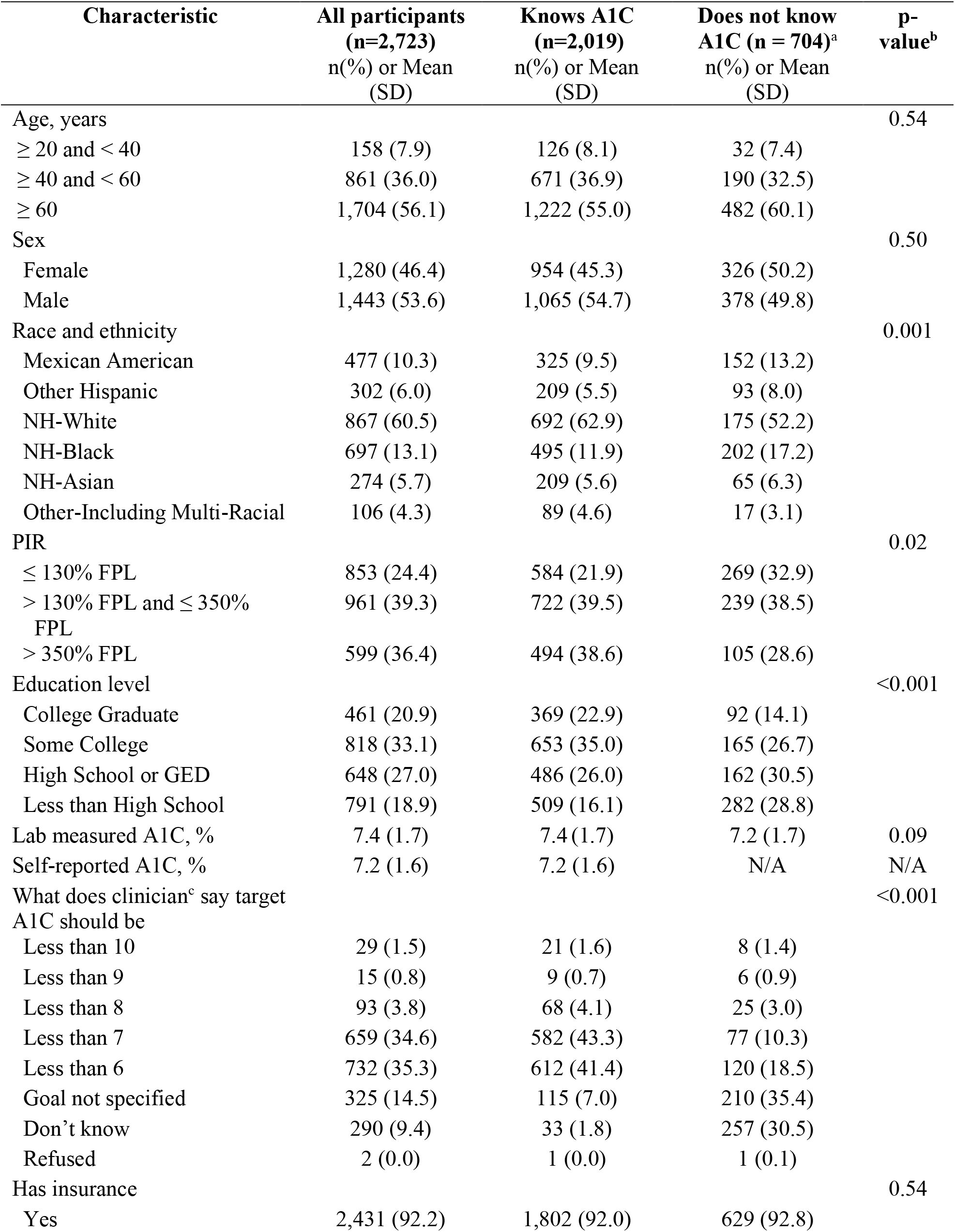
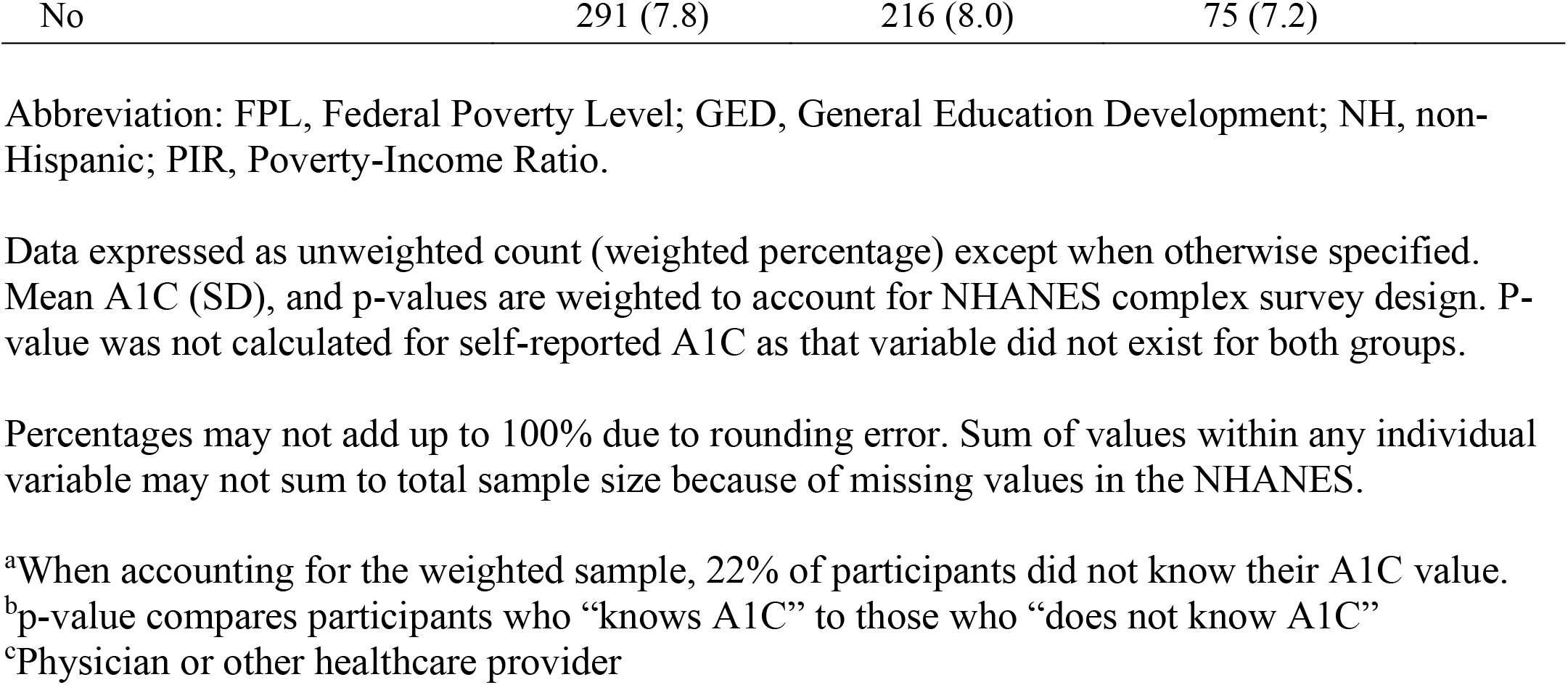
Characteristics of participants with self-reported diabetes by knowledge of A1C

| **Characteristic** | **All participants**<br>**(n=2,723)**<br>**n(%) or Mean**<br>**(SD)** | **Knows A1C**<br>**(n=2,019)**<br>**n(%) or Mean**<br>**(SD)** | **Does not know**<br>**A1C (n = 704)<sup>a</sup>**<br>**n(%) or Mean**<br>**(SD)** | **p-**<br>**value<sup>b</sup>** |
| --- | --- | --- | --- | --- |
| Age, years |  |  |  | 0.54 |
| ≥ 20 and < 40 | 158 (7.9) | 126 (8.1) | 32 (7.4) |  |
| ≥ 40 and < 60 | 861 (36.0) | 671 (36.9) | 190 (32.5) |  |
| ≥ 60 | 1,704 (56.1) | 1,222 (55.0) | 482 (60.1) |  |
| Sex |  |  |  | 0.50 |
| Female | 1,280 (46.4) | 954 (45.3) | 326 (50.2) |  |
| Male | 1,443 (53.6) | 1,065 (54.7) | 378 (49.8) |  |
| Race and ethnicity |  |  |  | 0.001 |
| Mexican American | 477 (10.3) | 325 (9.5) | 152 (13.2) |  |
| Other Hispanic | 302 (6.0) | 209 (5.5) | 93 (8.0) |  |
| NH-White | 867 (60.5) | 692 (62.9) | 175 (52.2) |  |
| NH-Black | 697 (13.1) | 495 (11.9) | 202 (17.2) |  |
| NH-Asian | 274 (5.7) | 209 (5.6) | 65 (6.3) |  |
| Other-Including Multi-Racial | 106 (4.3) | 89 (4.6) | 17 (3.1) |  |
| PIR |  |  |  | 0.02 |
| ≤ 130% FPL | 853 (24.4) | 584 (21.9) | 269 (32.9) |  |
| > 130% FPL and ≤ 350%<br>FPL | 961 (39.3) | 722 (39.5) | 239 (38.5) |  |
| > 350% FPL | 599 (36.4) | 494 (38.6) | 105 (28.6) |  |
| Education level |  |  |  | <0.001 |
| College Graduate | 461 (20.9) | 369 (22.9) | 92 (14.1) |  |
| Some College | 818 (33.1) | 653 (35.0) | 165 (26.7) |  |
| High School or GED | 648 (27.0) | 486 (26.0) | 162 (30.5) |  |
| Less than High School | 791 (18.9) | 509 (16.1) | 282 (28.8) |  |
| Lab measured A1C, % | 7.4 (1.7) | 7.4 (1.7) | 7.2 (1.7) | 0.09 |
| Self-reported A1C, % | 7.2 (1.6) | 7.2 (1.6) | N/A | N/A |
| What does clinician<sup>c</sup> say target<br>A1C should be |  |  |  | <0.001 |
| Less than 10 | 29 (1.5) | 21 (1.6) | 8 (1.4) |  |
| Less than 9 | 15 (0.8) | 9 (0.7) | 6 (0.9) |  |
| Less than 8 | 93 (3.8) | 68 (4.1) | 25 (3.0) |  |
| Less than 7 | 659 (34.6) | 582 (43.3) | 77 (10.3) |  |
| Less than 6 | 732 (35.3) | 612 (41.4) | 120 (18.5) |  |
| Goal not specified | 325 (14.5) | 115 (7.0) | 210 (35.4) |  |
| Don't know | 290 (9.4) | 33 (1.8) | 257 (30.5) |  |
| Refused | 2 (0.0) | 1 (0.0) | 1 (0.1) |  |
| Has insurance |  |  |  | 0.54 |
| Yes | 2,431 (92.2) | 1,802 (92.0) | 629 (92.8) |  |

|  |  |  |  |
| --- | --- | --- | --- |
| No | 291 (7.8) | 216 (8.0) | 75 (7.2) |

Abbreviation: FPL, Federal Poverty Level; GED, General Education Development; NH, non-Hispanic; PIR, Poverty-Income Ratio.
Data expressed as unweighted count (weighted percentage) except when otherwise specified. Mean A1C (SD), and p-values are weighted to account for NHANES complex survey design. P-value was not calculated for self-reported A1C as that variable did not exist for both groups.
Percentages may not add up to 100% due to rounding error. Sum of values within any individual variable may not sum to total sample size because of missing values in the NHANES.
<sup>a</sup>When accounting for the weighted sample, 22% of participants did not know their A1C value.
<sup>b</sup>p-value compares participants who “knows A1C” to those who “does not know A1C”
<sup>c</sup>Physician or other healthcare provider

Figure 2 presents a survey weighted bubble plot of self-reported A1C versus lab-measured A1C. The figure demonstrates that at around 8% A1C, the conditional mean of self-reported A1C was approximately the same as the conditional mean of lab-measured A1C. Above 9% A1C, the conditional mean of self-reported A1C was greater than the conditional mean of lab-measured A1C. Below 7% A1C, the conditional mean of self-reported A1C was lower than the conditional mean of lab-measured A1C. However, there was significant variation in these data, which can be visualized in the Bland and Altman plot (supplement figure 1). These data indicate a moderate correlation between self-reported and lab-measured A1C (r = 0.62, P < 0.001). The ICC indicated a moderate agreement between the two variables (0.58, 95% CI: 0.55-0.62, P < 0.001).

**Figure 2.**
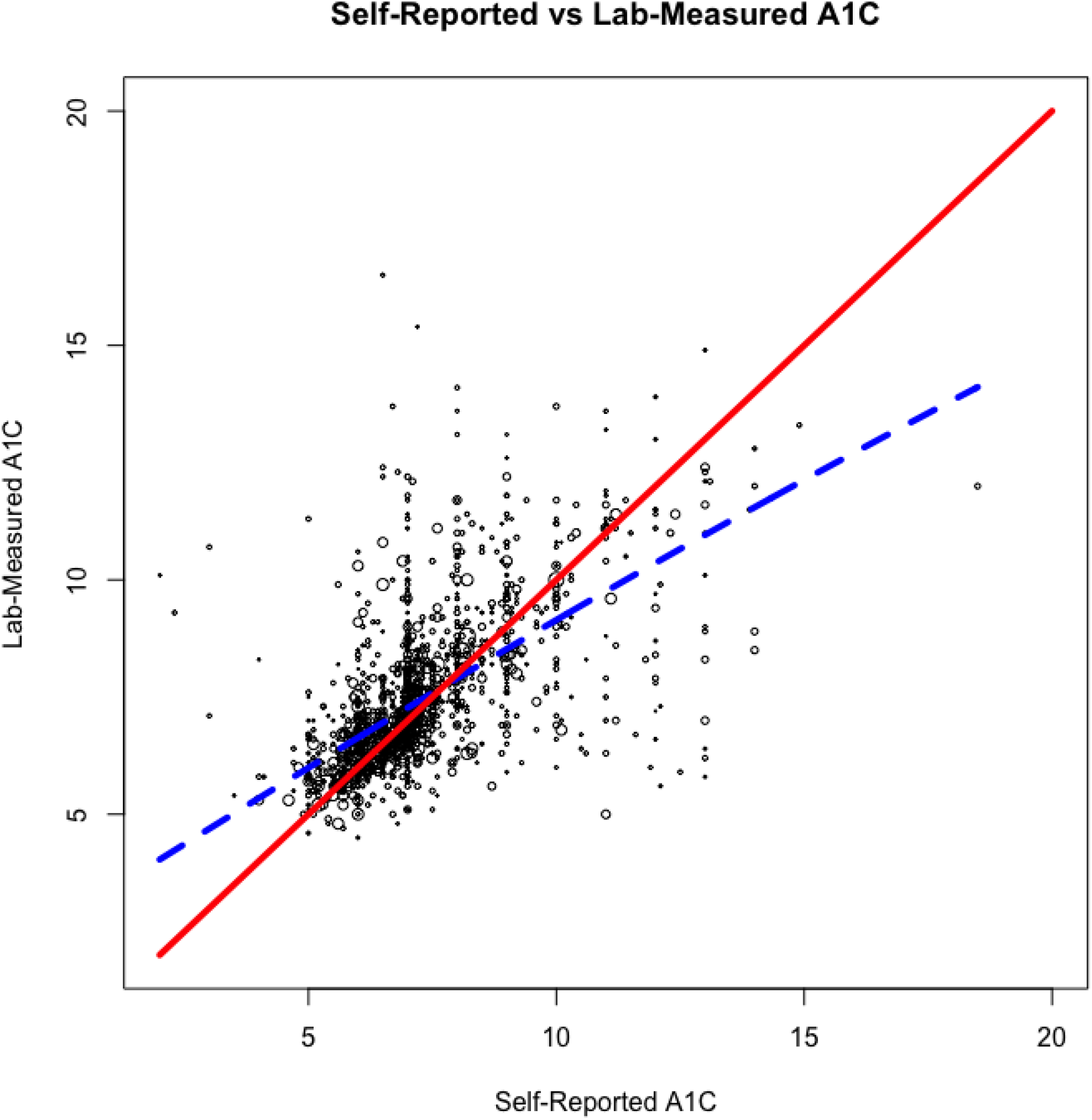
Bubble plot of self-reported A1C against lab-measured A1C. Size of bubbles indicates weight in survey. The solid line indicates a perfect correspondence between lab-measured and self-reported A1C, and the dashed line indicates the observed trend in our data.

In terms of Pearson correlations between self-reported and lab-measured A1C, college graduates had a notably lower correlation compared to participants with other educational attainment, at only 0.53. For participants without health insurance, the correlation was weak at only 0.45. Non-Hispanic Black and Non-Hispanic Asian participants had slightly lower correlations compared to other racial and ethnic groups, at 0.51 and 0.50 respectively. Participants with ages between 20 and 39, compared to older participants, had a slightly lower correlation at 0.55 (see table 2). Lin and Spearman correlation coefficients with 95% CIs were similar to Pearson correlations (see supplement table 1).

**Table 2.**
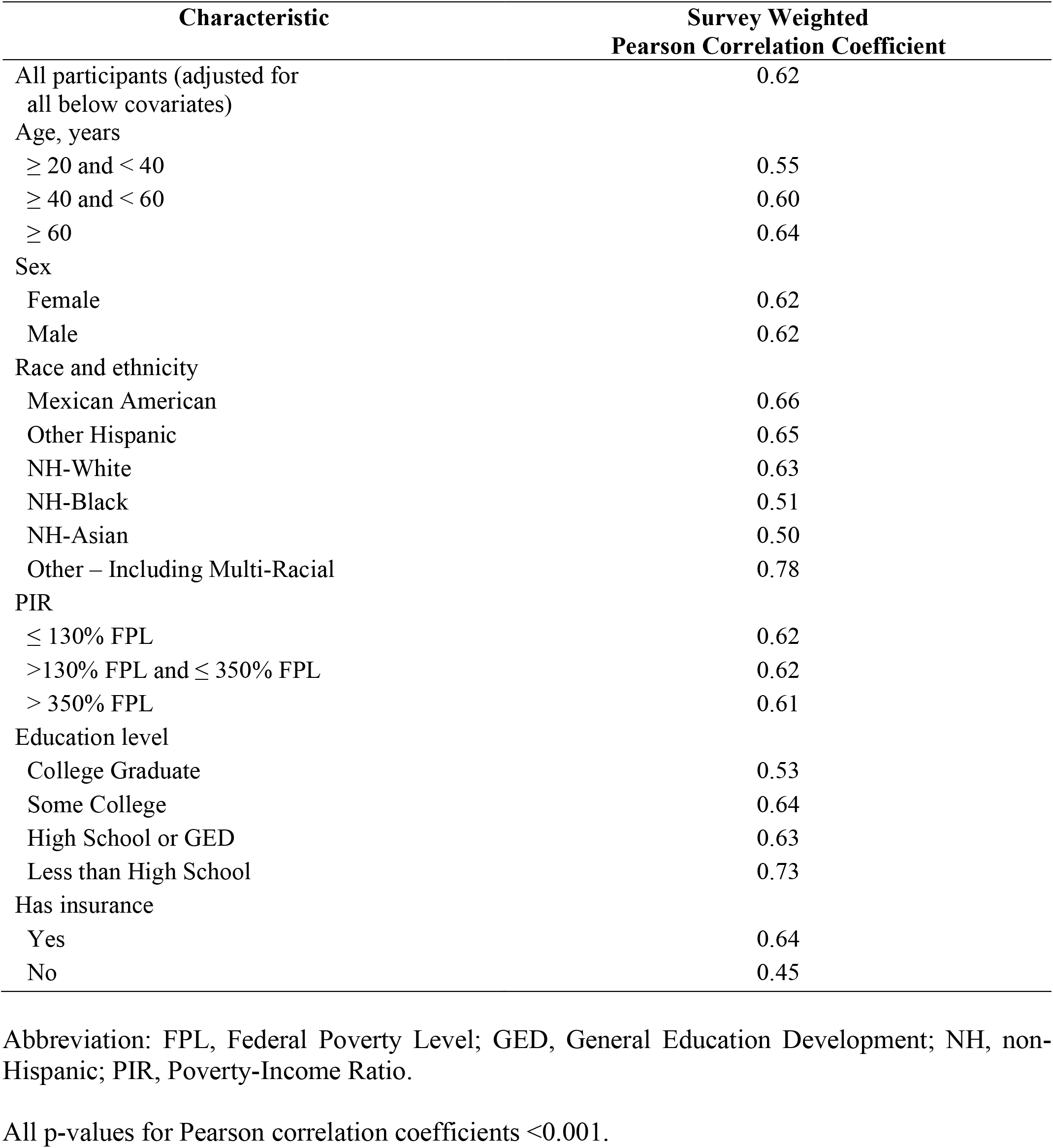
Correlations between self-reported and lab-measured A1C values among select subgroups NHANES 2013-2020

| Characteristic | Survey Weighted<br>Pearson Correlation Coefficient |
| --- | --- |
| All participants (adjusted for all below covariates) | 0.62 |
| Age, years |  |
| $\geq 20$ and $< 40$ | 0.55 |
| $\geq 40$ and $< 60$ | 0.60 |
| $\geq 60$ | 0.64 |
| Sex |  |
| Female | 0.62 |
| Male | 0.62 |
| Race and ethnicity |  |
| Mexican American | 0.66 |
| Other Hispanic | 0.65 |
| NH-White | 0.63 |
| NH-Black | 0.51 |
| NH-Asian | 0.50 |
| Other – Including Multi-Racial | 0.78 |
| PIR |  |
| $\leq 130\%$ FPL | 0.62 |
| $>130\%$ FPL and $\leq 350\%$ FPL | 0.62 |
| $> 350\%$ FPL | 0.61 |
| Education level |  |
| College Graduate | 0.53 |
| Some College | 0.64 |
| High School or GED | 0.63 |
| Less than High School | 0.73 |
| Has insurance |  |
| Yes | 0.64 |
| No | 0.45 |

Abbreviation: FPL, Federal Poverty Level; GED, General Education Development; NH, non-Hispanic; PIR, Poverty-Income Ratio.
All p-values for Pearson correlation coefficients $<0.001$ .

A survey-weighted confidence interval estimated that 52.0% (95% CI: 48.6%-55.0%) of participants who reported knowing their A1C were within 0.5% of their actual lab-measured value. Approximately 75.4% (95% CI: 72.4%-78.0%) of the participants had a difference of ≤ 1 between their self-reported and lab-measured A1C, while 9.5% (95% CI: 7.8%-12.0%) had self-reported A1C that was different by ≥ 2 compared to their lab-measured A1C. About 6% (95% CI: 4.6%-8.0%) of participants had self-reported A1C that differed by ≥ 3 from their lab-measured A1C.

In a logistic regression model to predict good glycemic control, defined as lab-measured A1C ≤ 7%, a 1-point increase in self-reported A1C corresponded to 0.87 (95% CI: 0.85-0.89) times lower odds of achieving glycemic control. It is important to note that the underlying correlation between self-reported and lab-measured A1C may contribute to these results. Compared to non-Hispanic White participants, non-Hispanic Black participants had 0.93 (95% CI: 0.87-0.99) and Mexican American participants had 0.89 (0.81-0.99) lower odds of good glycemic control. Non-Hispanic Asian and Other Hispanic participants had similar glycemic control to non-Hispanic White participants. Age, sex, and PIR were not statistically significant predictors of glycemic control in the model (see table 3).

**Table 3.** Odds of good (≤ 7%) lab-measured glycemic control against self-reported A1C and covariates

| Characteristic | Odds Ratio (95% CI) | p-value |
| --- | --- | --- |
| Self-reported A1C, % | 0.87 (0.85-0.89) | < 0.001 |
| Age (compared to $\geq 20$ and < 40) | | |
| $\geq 40$ and < 60 | 0.92 (0.78-1.08) | 0.31 |
| $\geq 60$ | 1.00 (0.86-1.17) | 0.98 |
| Sex (compared to Male) |  |  |
| Female | 1.04 (0.97-1.11) | 0.31 |
| Race and ethnicity (compared to NH-White) |  |  |
| Mexican American | 0.89 (0.81-0.99) | 0.03 |
| Other Hispanic | 0.97 (0.85-1.10) | 0.63 |
| NH-Black | 0.93 (0.87-0.99) | 0.03 |
| NH-Asian | 1.00 (0.90-1.11) | 0.96 |
| Other – Including Multi-Racial | 1.03 (0.91-1.18) | 0.62 |
| PIR (compared to $\leq 130\%$ FPL) | | |
| $>130\%$ FPL and $\leq 350\%$ FPL | 0.94 (0.85-1.04) | 0.24 |
| $> 350\%$ FPL | 0.96 (0.86-1.06) | 0.39 |
| Education level (compared to below High School) |  |  |
| College Graduate | 1.03 (0.92-1.17) | 0.56 |
| Some College | 1.03 (0.91-1.17) | 0.62 |
| High School or GED | 0.92 (0.82-1.03) | 0.13 |
| Has Insurance (compared to Yes) |  |  |
| No | 1.03 (0.88-1.21) | 0.68 |

Abbreviation: CI, confidence interval; FPL, Federal Poverty Level; GED, General Education Development; NH, non-Hispanic; PIR, Poverty-Income Ratio.

## 4. Discussion

Of the participants with self-reported diabetes and who indicated obtaining an A1C test at a healthcare setting within the last year, almost a quarter did not know the results from that test. This is consistent with findings from Memon and colleagues, which reported that 20% of participants did not know their last A1C test result [17]. However, Heisler and colleagues [19] and Trivedi and colleagues [15] found the number of participants who did not know their recent A1C result to be higher, ranging from 44% to 50%. The age of the participants in our study and the Memon [17] study were younger (mean of 60 and 56, respectively) compared to those in the study by Trivedi [15] (mean of 66). Unfortunately, the mean age of the participants in Heisler study was not provided [19]. The differences in the age of the participant may have contributed to the contrasting results. Additionally, our study found statistically significant differences by racial and ethnic backgrounds, PIR, and educational attainment among the individuals who did not know their A1C value compared to those who reported knowing it. Furthermore, participants with diabetes who did not know their A1C value were more likely to not know or have an unspecified clinician-provided target A1C goal.

Self-reported A1C had a moderate correlation with lab-measured A1C. About half of patients with diabetes had A1C concordance – where their self-reported A1C differed from lab-measured A1C by ≤ 0.5%. The degree of concordance in our study was lower than that reported by Trivedi [15] (52.0% vs. 78.3%) which used the same operational definition of concordance. Participants in the Trivedi study were from eight European countries and little sociodemographic information, including race and ethnicity, was provided [15]. Thus, it is not possible to examine if individual sociodemographic characteristics may be contributing to the difference in findings. In a logistic regression model, self-reported A1C was a statistically significant predictor of glycemic control with a 1-point increase in self-reported A1C corresponding to 0.87 (95% CI: 0.85-0.89) times lower odds of achieving glycemic control.

Our study found that self-reported and lab-measured A1C were moderately correlated. However, individuals with higher A1C levels (≥ 9%) tended to overestimate the lab-measured value and individuals with lower A1C levels (< 7%) tended to underestimate the lab-measured value. These findings differ from what has been previously reported. Trivedi and colleagues in an international cohort found self-reported and lab-measured A1C to be highly correlated [15]. Fowles and colleagues found that self-reported A1C tended to be overestimated compared to lab-measured A1C [45]. Whereas, Memon and colleagues, found that self-reported A1C tended to be closer to the ideal 7% A1C value compared to lab-measures [17]. Given the overall lack of consistent findings regarding the concordance of self-reported and lab-measured A1C, studies that have relied only on self-reported A1C should be interpreted with caution. Furthermore, future studies should prioritize lab-measured A1C, especially if examining interventions aimed at improving glycemic control and diabetes management.

The substantial portion of patients with diabetes who did not know their A1C reinforces the need for improved patient-provider communication about A1C. Patient understanding of A1C is crucial to improving diabetes self-management techniques. Recent studies have indicated that patients with better knowledge of their A1C have better overall care, diabetes management, medication knowledge, adherence, and have increased treatment satisfaction [46-48]. Aung and colleagues have elucidated that self-knowledge of A1C is additionally associated with patients taking a more active role in their healthcare [49]. Prior research has not focused on changes in patient knowledge of their own A1C value, and rather has focused on improvements in A1C measured in a healthcare setting.

Strengths of this study include the generalizability of NHANES to the U.S. population, NHANES oversampling methods, and the standardization of NHANES protocols. Potential limitations include a relatively small sample of participants with diabetes in the study, though theoretically proportional to the U.S. population. NHANES did not allow us to differentiate between patients diagnosed with Type 1 and Type 2 diabetes. As NHANES only asked for A1C tests in a healthcare setting among patients with diagnosed diabetes, we can neither make conclusions about patients with diabetes who have self-tested for their A1C nor for those who have undiagnosed diabetes. Additionally, as A1C is a measure of glycemic control, the moderate concordance between self-reported and lab-measured A1C could be due to changes in glycemic control since the last A1C test rather than inaccurate self-reporting. Furthermore, as the data were self-reported, social desirability bias may apply, causing underreporting of high A1C values. As NHANES is cross-sectional, we cannot make any assertions about causality and can only measure associations.

## 5. Conclusion

Despite receiving an A1C test, many patients report not knowing the results. Sociodemographic characteristics associated with not knowing one’s A1C values were identifying as a racial or ethnic group other than White, having a lower PIR, and having a less formal education. These findings suggest that certain groups may be receiving inadequate diabetes care and education, which could be negatively affecting their diabetes self-management. Among those who knew their previous A1C value, nearly half over- or under-estimated their A1C by > 0.5%. Assessing how A1C results are communicated and improving patients’ knowledge and understanding about A1C should be priorities for providers to maximize diabetes self-management and to mitigate the long-term negative consequences of poor glycemic control.

## Data Availability

All data used in this study can be obtained at the NHANES website. Only NHANES data were used in this study. Upon contacting the authors, the files used for analyses can be provided.

## Acknowledgements

None

## References

[1] Centers for Disease Control and Prevention. National Diabetes Statistics Report website. https://www.cdc.gov/diabetes/data/statistics-report/index.html, (Accessed March 21, 2022).

[2] American Diabetes Association, Economic Costs of Diabetes in the U.S. in 2017, Diabetes Care 41 (5) (2018) 917–928, https://doi.org/10.2337/dci18-0007.

[3] Menke, S. Casagrande, L. Geiss, C.C. Cowie, Prevalence of and Trends in Diabetes Among Adults in the United States, 1988–2012, JAMA 314 (10) (2015) 1021–1029, https://doi.org/10.1001/jama.2015.10029.

[4] M. Fang, D. Wang, J. Coresh, E. Selvin, Trends in Diabetes Treatment and Control in U.S. Adults, 1999–2018, N Engl J Med 384 (23) (2021) 2219–2228, https://doi.org/10.1056/NEJMsa2032271.

[5] N. Papadopoulou-Marketou, G.P. Chrousos, C. Kanaka-Gantenbein, Diabetic nephropathy in type 1 diabetes: a review of early natural history, pathogenesis, and diagnosis, Diabetes Metab Res Rev. 33 (2) (2017), https://doi.org/10.1002/dmrr.2841.

[6] C.G. Jolivalt, K.E. Frizzi, L. Guernsey, et al., Peripheral Neuropathy in Mouse Models of Diabetes, Curr Protoc Mouse Biol. 6 (3) (2016) 223–255, https://doi.org/10.1002/cpmo.11.

[7] H.P. Hammes, K.D. Lemmen, B. Bertram, Diabetic Retinopathy and Maculopathy, Exp Clin Endocrinol Diabetes 129 (S 01) (2021) S64–s69, https://doi.org/10.1055/a-1284-6223.

[8] Modi, A. Agrawal, F. Morgan, Euglycemic Diabetic Ketoacidosis: A Review, Curr Diabetes Rev. 13 (3) (2017) 315–321, https://doi.org/10.2174/1573399812666160421121307.

[9] D. Sun, T. Zhou, Y. Heianza, et al., Type 2 Diabetes and Hypertension, Circ Res. 124 (6) (2019) 930–937, https://doi.org/10.1161/CIRCRESAHA.118.314487.

[10] D.M. Nathan, S. Genuth, J. Lachin, et al., The effect of intensive treatment of diabetes on the development and progression of long-term complications in insulin-dependent diabetes mellitus, N Engl J Med 329 (14) (1993) 977–86, https://doi.org/10.1056/nejm199309303291401.

[11] D.M. Nathan, D.E. Singer, K. Hurxthal, J.D. Goodson, The clinical information value of the glycosylated hemoglobin assay, N Engl J Med 310 (6) (1984) 341–6. https://doi.org/10.1056/nejm198402093100602.

[12] S.I. Sherwani, H.A. Khan, A. Ekhzaimy, A. Masood, M.K. Sakharkar, Significance of HbA1c Test in Diagnosis and Prognosis of Diabetic Patients, Biomark Insights 11 (2016) 95–104, https://doi.org/10.4137/BMI.S38440.

[13] American Diabetes Association. Understanding A1C: A1C does it all. https://www.diabetes.org/diabetes/a1c, (Accessed Mar 5, 2022).

[14] A.M. Delamater, Clinical Use of Hemoglobin A1c to Improve Diabetes Management, Clinical Diabetes 24 (1) (2006) 6–8, https://doi.org/10.2337/diaclin.24.1.6

[15] H. Trivedi, L.J. Gray, S. Seidu S, et al., Self-knowledge of HbA1c in people with Type 2 Diabetes Mellitus and its association with glycaemic control, Prim Care Diabetes 11 (5) (2017) 414–420, https://doi.org/10.1016/j.pcd.2017.03.011.

[16] J.P. Twarog, A.M. Charyalu, M.R. Subhani, P. Shrestha, E. Peraj, Differences in HbA1C% screening among U.S. adults diagnosed with diabetes: Findings from the National Health and Nutrition Examination Survey (NHANES), Prim Care Diabetes 12 (6) (2018) 533–536, https://doi.org/10.1016/j.pcd.2018.07.006.

[17] R. Memon, D. Levitt, S.R. Salgado Nunez Del Prado, et al., Knowledge of Hemoglobin A1c and Glycemic Control in an Urban Population, Cureus 13 (3) (2021) e13995, https://doi.org/10.7759/cureus.13995.

[18] Gopalan, K. Kellom, K. McDonough, M.M. Schapira, Exploring how patients understand and assess their diabetes control, BMC Endocr Disord 18 (1) (2018) 79, https://doi.org/10.1186/s12902-018-0309-4.

[19] M. Heisler, J.D. Piette, M. Spencer, E. Kieffer, S. Vijan, The relationship between knowledge of recent HbA1c values and diabetes care understanding and self-management, Diabetes Care 28 (4) (2005) 816–822, https://doi.org/10.2337/diacare.28.4.816.

[20] G. Zipf, M. Chiappa, K.S. Porter, Y. Ostchega, B.G. Lewis, J. Dostal, National Health and Nutrition Examination Survey: Plan and operations, 1999–2010. National Center for Health Statistics. Vital Health Stat 1 (56) (2013), https://www.cdc.gov/nchs/data/series/sr_01/sr01_056.pdf (Accessed Feb 26, 2023)

[21] National Center for Health Statistics (U.S.). About the National Health and Nutrition Examination Survey, Updated September 15, 2017, https://www.cdc.gov/nchs/nhanes/about_nhanes.htm (Accessed January 20, 2022).

[22] National Center for Health Statistics. NHANES Questionnaires, Datasets, and Related Documentation, https://www.n.cdc.gov/nchs/nhanes/Default.aspx (Accessed March 9, 2022).

[23] C.R. Mottishaw, S. Becker, B. Smith, et al., Insights into the Progression of Labile Hb A(1c) to Stable Hb A(1c) via a Mechanistic Assessment of 2,3-Bisphosphoglycerate Facilitation of the Slow Nonenzymatic Glycation Process, Hemoglobin. Jan 43 (1) (2019) 42–49, https://doi.org/10.1080/03630269.2019.1597731

[24] National Health and Nutrition Examination Survey. Glycohemoglobin (P_GHB), https://www.n.cdc.gov/Nchs/Nhanes/2017-2018/P_GHB.htm, (Accessed March 13, 2022).

[25] National Health and Nutrition Examination Survey. Glycohemoglobin (GHB_I), https://www.n.cdc.gov/Nchs/Nhanes/2015-2016/GHB_I.htm, (Accessed March 14, 2022).

[26] National Health and Nutrition Examination Survey. Glycohemoglobin (GHB_H), https://www.n.cdc.gov/Nchs/Nhanes/2013-2014/GHB_H.htm (Accessed March 15, 2022).

[27] G. Kaiafa, S. Veneti, G. Polychronopoulos, D. Pilalas, et al., Is HbA1c an ideal biomarker of well-controlled diabetes?, Postgrad Med J 97 (1148) (2021) 380–383, https://doi.org/10.1136/postgradmedj-2020-138756.

[28] F. Guo, W.T. Garvey, Trends in Cardiovascular Health Metrics in Obese Adults: National Health and Nutrition Examination Survey (NHANES), 1988–2014, J Am Heart Assoc 5 (7) (2016), https://doi.org/10.1161/jaha.116.003619.

[29] C.L. Ogden, T.H. Fakhouri, M.D. Carroll, et al., Prevalence of Obesity Among Adults, by Household Income and Education - United States, 2011–2014, MMWR Morb Mortal Wkly Rep 66 (50) (2017) 1369–1373, https://doi.org/10.15585/mmwr.mm6650a1.

[30] S.L. Jackson, E.C. Yang, Z. Zhang, Income Disparities and Cardiovascular Risk Factors Among Adolescents, Pediatrics 142 (5) (2018) e20181089, https://doi.org/10.1542/peds.2018-1089.

[31] National Health and Nutrition Examination Survey. Diabetes (DIQ_I), https://www.n.cdc.gov/Nchs/Nhanes/2015-2016/DIQ_I.htm#DIQ010, (Accessed March 19, 2022).

[32] R: A Language and Environment for Statistical Computing. 2021. https://www.R-project.org/ (Accessed March 25, 2022)

[33] T. Lumley, Analysis of Complex Survey Samples. J Stat Softw. 9 (1) (2004) 1–19. https://doi.org/10.18637/jss.v009.i08

[34] survey: Analysis of Complex Survey Samples, http://r-survey.r-forge.r-project.org/survey/ (Accessed March 25, 2022).

[35] jtools: Analysis and Presentation of Social Scientific Data, https://jtools.jacob-long.com (Accessed March 25, 2022).

[36] Gamer M, Lemon J, Singh IFP. irr: Various Coefficients of Interrater Reliability and Agreement, https://cran.r-project.org/web/packages/irr/index.html (Accessed Feb 13, 2022).

[37] C.L. Johnson, R. Paulose-Ram, C.L. Ogden, et al. National Health and Nutrition Examination Survey: Analytic guidelines, 1999–2010. National Center for Health Statistics. Vital Health Stat 2 (161) (2013), https://www.cdc.gov/nchs/data/series/sr_02/sr02_161.pdf (Accessed Feb 20, 2023).

[38] S.Y. Chen, Z. Feng, X. Yi, A general introduction to adjustment for multiple comparisons, J Thorac Dis. 9 (6) (2017) 1725–1729, https://doi.org/10.21037/jtd.2017.05.34.

[39] J.M. Bland, D.G. Altman, A note on the use of the intraclass correlation coefficient in the evaluation of agreement between two methods of measurement, Comput Biol Med 20 (5) (1990) 337–40, https://doi.org/10.1016/0010-4825(90)90013-f

[40] K.O. McGraw, S.P. Wong, Forming inferences about some intraclass correlation coefficients, Psychological Methods 1 (19960 30–46, https://doi.org/10.1037/1082-989X.1.1.30

[41] T.K. Koo, M.Y. Li, A Guideline of Selecting and Reporting Intraclass Correlation Coefficients for Reliability Research, J Chiropr Med 15 (2) (2016) 155–63, https://doi.org/10.1016/j.jcm.2016.02.012

[42] S. Chalew, R. Gomez, A. Vargas, et al., Hemoglobin A1c, frequency of glucose testing and social disadvantage: Metrics of racial health disparity in youth with type 1 diabetes, Journal of Diabetes and its Complications, 32(12) (2018) 1085–1090, https://doi.org/10.1016/j.jdiacomp.2018.02.008

[43] M. Mongraw-Chaffin, A.G. Bertoni, S.H. Golden, et al., Association of Low Fasting Glucose and HbA1c With Cardiovascular Disease and Mortality: The MESA Study, J Endocr Soc. 3 (5) (2019) 892–901, https://doi.org/10.1210/js.2019-00033

[44] T.S. King, V.M. Chinchilli. Robust estimators of the concordance correlation coefficient. J Biopharm Stat. 2001;11(3):83–105. doi:10.1081/BIP-100107651

[45] J.B. Fowles, K. Rosheim, E.J. Fowler, C. Craft, L. Arrichiello, The validity of self-reported diabetes quality of care measures, Int J Qual Health Care 11 (5) (1999) 407–12, https://doi.org/10.1093/intqhc/11.5.407.

[46] S. Yang, W. Kong, C. Hsue, et al., Knowledge of A1c Predicts Diabetes Self-Management and A1c Level among Chinese Patients with Type 2 Diabetes, PLoS One 11 (3) (2016) e0150753, https://doi.org/10.1371/journal.pone.0150753.

[47] R. Alsaedi, K. McKeirnan, Literature Review of Type 2 Diabetes Management and Health Literacy, Diabetes Spectr 34 (4) (2021) 399–406, https://doi.org/10.2337/ds21-0014

[48] N Iqbal, C. Morgan, H. Maksoud, I. Idris, Improving patients’ knowledge on the relationship between HbA1c and mean plasma glucose improves glycaemic control among persons with poorly controlled diabetes, Ann Clin Biochem 45 (Pt 5) (2008) 504–7, https://doi.org/10.1258/acb.2008.008034.

[49] E. Aung, M. Donald, G.M. Williams, J.R. Coll, S.A. Doi, Joint influence of Patient-Assessed Chronic Illness Care and patient activation on glycaemic control in type 2 diabetes, Int J Qual Health Care 27 (2) (2015) 117–24, http://doi.org/10.1093/intqhc/mzv001

