## Supplemental Figure 1 for "Discordance Between Self-Reported and Lab-Measured A1C Among U.S. Adults with Diabetes: Findings from the National Health and Nutrition Examination Survey (2013-2020)"

**Supplement Figure 1.** Bland and Altman Plot of Self-reported and Lab-measured A1C. The mean difference between the two measures was -0.22 (95% CI: -3.16, 2.72). The plot indicates a moderate agreement but high variation between these two variables.

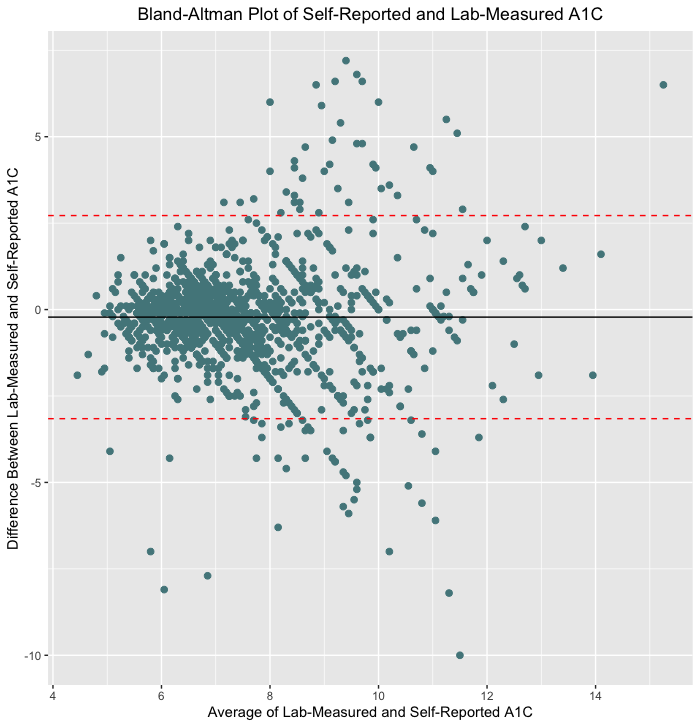
