## Supplemental Table 1 for "Discordance Between Self-Reported and Lab-Measured A1C Among U.S. Adults with Diabetes: Findings from the National Health and Nutrition Examination Survey (2013-2020)"

**Supplement Table 1.** Lin and Spearman Correlations between self-reported and lab-measured A1C values among select subgroups NHANES 2013-2020

| **Characteristic** | **Lin’s Correlation Coefficient,**  $\boldsymbol{\rho}_{\boldsymbol{c}}$ **(95% CI)** | **Spearman Correlation Coefficient,**  $\boldsymbol{\rho}$ **(95% CI)** |
| --- | --- | --- |
| Age, years |  |  |
| ≥ 20 and < 40 | 0.55 (0.35-0.72) | 0.61 (0.43-0.75) |
| ≥ 40 and <60 | 0.60 (0.51-0.68) | 0.66 (0.59-0.72) |
| ≥ 60 | 0.63 (0.57-0.70) | 0.68 (0.63-0.72) |
| Sex |  |  |
| Female | 0.62 (0.55-0.69) | 0.72 (0.66-0.76) |
| Male | 0.62 (0.54-0.69) | 0.63 (0.57-0.68) |
| Race and ethnicity |  |  |
| Mexican American | 0.62 (0.48-0.74) | 0.64 (0.52-0.74) |
| Other Hispanic | 0.64 (0.49-0.76) | 0.74 (0.61-0.83) |
| NH White | 0.63 (0.55-0.70) | 0.68 (0.62-0.73) |
| NH Black | 0.49 (0.37-0.60) | 0.61 (0.52-0.68) |
| NH Asian | 0.50 (0.32-0.66) | 0.61 (0.48-0.74) |
| Other – Including Multi-Racial | 0.78 (0.59-0.88) | 0.75 (0.59-0.85) |
| PIR |  |  |
| ≤ 130% FPL | 0.62 (0.52-0.70) | 0.67 (0.59-0.74) |
| >130% FPL and ≤ 350% FPL | 0.61 (0.52-0.69) | 0.65 (0.58-0.70) |
| > 350% FPL | 0.60 (0.50-0.68) | 0.67 (0.61-0.74) |
| Education level |  |  |
| College Graduate | 0.53 (0.39-0.65) | 0.57 (0.47-0.66) |
| Some College | 0.64 (0.56-0.71) | 0.74 (0.69-0.79) |
| High School or GED | 0.60 (0.49-0.71) | 0.66 (0.59-0.73) |
| Less than High School | 0.73 (0.64-0.80) | 0.72 (0.64-0.78) |
| Has insurance |  |  |
| Yes | 0.63 (0.58-0.68) | 0.68 (0.65-0.72) |
| No | 0.45 (0.25-0.62) | 0.50 (0.32-0.66) |

Abbreviation: CI, confidence interval; FPL, Federal Poverty Level; GED, General Education Development; NH, non-Hispanic; PIR, Poverty-Income Ratio.
